# EBUS Diagnostic Yield for Sarcoidosis in Hilar vs. Mediastinal Lymph Nodes

**DOI:** 10.1101/2025.04.20.25326129

**Authors:** Naeman Mahmood, Steven Wolf, Raj Dash, Xiaofei Wang, Scott L. Shofer, Coral X. Giovacchini, Michael Dorry, Hakim Azfar Ali, Kamran Mahmood, Momen M. Wahidi

## Abstract

**Background:** Pulmonary sarcoidosis is diagnosed by endobronchial ultrasound-guided transbronchial needle aspirate (EBUS-TBNA) of hilar and mediastinal lymph nodes and the finding of non-caseating granulomatous inflammation. There are currently no guidelines about which lymph node stations to sample to optimize the diagnostic yield and it is unclear if there is a difference in the yield between hilar and mediastinal lymph node stations.

**Methods:** A retrospective study was performed to assess the difference in the diagnostic yield of EBUS-TBNA for non-caseating granulomas between hilar and mediastinal lymph nodes.

**Results:** Two hundred twenty-five patients with suspicion of sarcoidosis underwent EBUS-TBNA for evaluation of hilar and mediastinal lymphadenopathy. The yield of EBUS-TBNA for non-caseating granulomas was 61.8% vs. 65.5%, P = 0.46, for hilar and mediastinal lymph nodes, respectively. The sensitivity for sarcoidosis of EBUS-TBNA of hilar vs. mediastinal nodes was 66.9% (95% confidence interval or CI, 58.9%-74.9%) vs. 71.1% (95% CI, 65.3%-76.9%). The specificity for sarcoidosis of EBUS-TBNA of both hilar and mediastinal nodes was 100%. The diagnostic yield for non-caseating granulomas in patients who underwent hilar nodes biopsy only, mediastinal nodes biopsy only, and both hilar and mediastinal nodes biopsy was 71.4%, 67%, and 73.1%, respectively (P=0.63). In multivariable logistic regression analysis, the diagnostic yield of EBUS-TBNA was only associated with age (OR 0.96; 95% CI 0.94-0.98; P <0.01).

**Conclusions:** The yield of EBUS-TBNA for non-caseating granulomas in patients with suspected sarcoidosis was similar between the hilar and mediastinal lymph nodes.

## Introduction

Sarcoidosis is a systemic disease of unclear etiology that is characterized by non-caseating granulomatous inflammation which can affect any organ (1). The estimated prevalence of sarcoidosis in the United States is about 60 per 100,000 people, with a higher prevalence in African Americans (2–4). Pulmonary involvement is seen in about 90% of sarcoidosis patients and often presents with hilar and mediastinal lymphadenopathy and a variety of pulmonary parenchymal findings on radiographic studies (5, 6). Pulmonary sarcoidosis is diagnosed based on the clinical presentation and radiological findings, and a biopsy is often required to confirm the diagnosis (6, 7). The standard targets of the biopsy are hilar and mediastinal lymph nodes in patients with lymphadenopathy seen in stage I and II pulmonary sarcoidosis (1, 6).

Endobronchial ultrasound guided transbronchial needle aspiration (EBUS-TBNA) has become the procedure of choice, as the diagnostic yield is better than conventional TBNA and it is less invasive compared to mediastinoscopy (1, 7–10). The current guidelines also endorse EBUS-TBNA for the evaluation of sarcoidosis (7, 11, 12). However, it remains unknown if the diagnostic yield of EBUS-TBNA for non-caseating granulomatous inflammation varies between hilar and mediastinal nodal stations. Compared to the hilar node, the mediastinal nodes are usually larger and easier to access, and if there is a difference in the diagnostic yield, it can inform the diagnostic approach. We conducted this study to compare the diagnostic yield of EBUS-TBNA for non-caseating granulomatous inflammation between the hilar and mediastinal nodes. We present this manuscript in accordance with the STARD reporting checklist.

## Methods

We retrospectively reviewed the medical records of all patients who underwent EBUS-TBNA at a single tertiary care hospital to investigate hilar and mediastinal lymphadenopathy seen on the CT chest for suspected sarcoidosis between January 2015 and March 2021. The diagnostic yield of EBUS-TBNA for non-caseating granulomas was defined as the percentage of lymph nodes or patients with cytological confirmation of non-caseating granulomas over a total number of nodes or patients biopsied, respectively. This definition has been previously used in other studies (1, 8). The non-caseating granulomas were identified when groups of closely aggregated epithelioid histiocytes with ovoid or elongated nuclei without necrosis were seen, often with multinucleated giant cells, and negative acid fast bacilli and fungal stains or cultures (13). The data collection included demographics and details of the procedure, including the lymph node stations biopsied, lymph node size, biopsy needle size, number of biopsy passes, and the lymph node biopsy results. The hilar and mediastinal nodal stations were identified using the IASLC lymph node map and suggested landmarks (14). EBUS accessible mediastinal stations included stations 2, 3P, 4, and 7, and hilar stations included 10, 11, and 12. Definitive sarcoidosis was defined based on typical clinical and radiographic features and the finding of non-caseating granulomas on EBUS-TBNA, transbronchial biopsy, endobronchial biopsy, mediastinoscopy, or other organ biopsies on a one-year follow-up (7, 8). Probable sarcoidosis was defined as clinical and radiographic features of sarcoidosis and the absence of alternate diagnosis when the biopsies were negative, but the treating physicians diagnosed the patient with sarcoidosis on a one-year follow-up (7, 8).

EBUS was performed under moderate sedation until February 2016 and with general anesthesia afterwards. The EBUS bronchoscope (Olympus, Center Valley, PA, USA) was introduced through the airway, and target nodes were identified. The EBUS needle (Olympus) was introduced into the target lesion under direct ultrasound visualization, the stylet was removed, and suction was applied. The needle was agitated to collect the specimen, the suction was turned off, the needle was removed, and the aspirate was further processed.

One to two aspirates were placed on slides and stained with Diff Quik for rapid on-site cytological evaluation (ROSE) to determine specimen adequacy.(15) The rest of the passes were placed in 95% formalin or sterile saline for cytological examination or cultures, per the standard institutional work flow as previously described.(16)

The authors are accountable for all aspects of the work in ensuring that questions related to the accuracy or integrity of any part of the work are appropriately investigated and resolved. The study was conducted in accordance with the Declaration of Helsinki (as revised in 2013). The study was approved by the institutional review board of Duke University (IRB protocol Pro00107694) and individual consent for this retrospective analysis was waived.

### Statistical Analyses

Continuous variables were summarized as median and interquartile range (first quartile, Q1-third quartile, Q3) and categorical data as percentages. Continuous outcomes were compared with a Wilcoxon rank sum test, and categorical outcomes with a chi-square test. A generalized estimating equation (GEE) model for the binomial distribution with a logit link was fit to evaluate the relationship of the binary outcome diagnostic yield and baseline variables. This model also accounted for the repeated measures taken by the same patient. An exchangeable correlation structure was selected using the QIC model selection method. We excluded certain variables from the adjusted model due to a lack of association with the outcome. However, we made an exception for the hilar vs. mediastinal lymph node and needle size due to their clinical relevance to the outcome. A two-sided P-value of ≤0.05 was considered statistically significant.

## Results

The study included two hundred twenty-five patients, and their characteristics are shown in Table 1. The median age (interquartile range, IQR) was 54 (43–63) years. Females accounted for 61.3% of the cohort. There were 44.9% African Americans, 48.4% Caucasians, 1.8% Asians, and others. There were 4.9% smokers and 2.7% patients who used vaping devices.

**Table 1.** Demographics and characteristics of patients.

| Characteristics | Patients (N=225) |
| --- | --- |
| Age, median (IQR) years | 54 (43-63) |
| Gender- female, n (%) | 138 (61.3) |
| Race– n (%) |  |
| African-American | 101 (44.9) |
| Caucasian | 109 (48.4) |
| Asian | 4 (1.8) |
| American Indian/Alaskan | 1 (0.4) |
| Native | 2 (0.9) |
| Unavailable | 8 (3.6) |
| BMI- median (IQR), kg/m<sup>2</sup> | 30.7 (26.8-35.1) |
| Smoking Status– n (%) |  |
| Non-smoker | 214 (95.1) |
| Smoker | 11 (4.9) |
| Vaping Status- n (%) |  |
| No | 219 (97.3) |
| Yes | 6 (2.7) |

The EBUS-TBNA was performed on 144 hilar and 255 mediastinal nodes (**Table 2**). The median lymph node size on EBUS for hilar and mediastinal nodes was 12 mm and 18 mm, respectively (P <0.0001). The needle sizes included 19G, 21G, and 22G for biopsy of 63.2%, 4.2%, and 26.4% hilar and 62%, 3.5%, and 26.3% mediastinal nodes, respectively (P=0.90). Median (IQR) biopsy passes were 4 (3–5) for hilar and 4 (4–5) for hilar and mediastinal nodes, respectively (P=0.03). The final cytology results from EBUS-TBNA showed non-caseating granulomas, normal lymphocytes, cancer, and blood or bronchial epithelium in 61.8%, 24.3%, 0, and 3.9% hilar, and 65.5%, 23.5%, 0.8%, and 10.2% mediastinal nodes, respectively (P=0.49). No complications were reported from the procedures.

**Table 2:** EBUS characteristics and results.

|  | Hilar Nodes<br>N=144 | Mediastinal<br>Nodes<br>N=255 | P-value for<br>difference |
| --- | --- | --- | --- |
| Lymph node stations- n (%) |  |  | <0.0001* |
| 2R | 0 | 2 (0.8) |  |
| 2L | 0 | 1 (0.4) |  |
| 3P | 0 | 1 (0.4) |  |
| 4R | 0 | 62 (24.3) |  |
| 4L | 0 | 13 (5.1) |  |
| 7 | 0 | 176 (69) |  |
| 10R | 2 (1.4) | 0 |  |
| 10L | 0 | 0 |  |
| 11R | 62 (43.1) | 0 |  |
| 11L | 74 (51.4) | 0 |  |
| 12R | 6 (4.2) | 0 |  |
| 12L | 0 | 0 |  |
| Lymph node size on EBUS in mm,<br>median (IQR) | 12 (10-16) | 18 (11-23) | <0.0001** |
| Biopsy needle size- n (%) |  |  | 0.90* |
| 19G | 91 (63.2) | 158 (62) |  |
| 21G | 6 (4.2) | 9 (3.5) |  |
| 22G | 38 (26.4) | 67 (26.3) |  |
| Not available | 9 (6.3) | 21 (8.2) |  |
| Biopsy passes, median (IQR) | 4 (3-5) | 4 (4-5) | 0.03** |
| Lymph node cytology results, n (%) |  |  | 0.49* |
| Non-caseating granulomas | 89 (61.8) | 167 (65.5) |  |
| Normal lymphocytes | 35 (24.3) | 60 (23.5) |  |
| Cancer | 0 | 2 (0.8) |  |
| Blood or bronchial epithelial cells | 20 (13.9) | 26 (10.2) |  |

\*Chi-Square P-value
\*\*Wilcoxon Rank Sum Test P-value

As shown in **Table 3**, the diagnostic yield of EBUS-TBNA for non-caseating granulomas was 61.8% for hilar nodes and 65.5% for mediastinal nodes (P =0.46). The diagnostic yield for non-caseating granulomas in patients who underwent hilar nodes biopsy only, mediastinal nodes biopsy only, and both hilar and mediastinal nodes biopsy was 71.4%, 67%, and 73.1%, respectively (P=0.63), as shown in **Table 4** and **Figure 1**. Non-caseating granulomas were seen with transbronchial biopsies in 13.8% of patients and endobronchial biopsies in 12.9% of patients, as shown in Supplementary Material **Table S1**.

**Figure 1:**
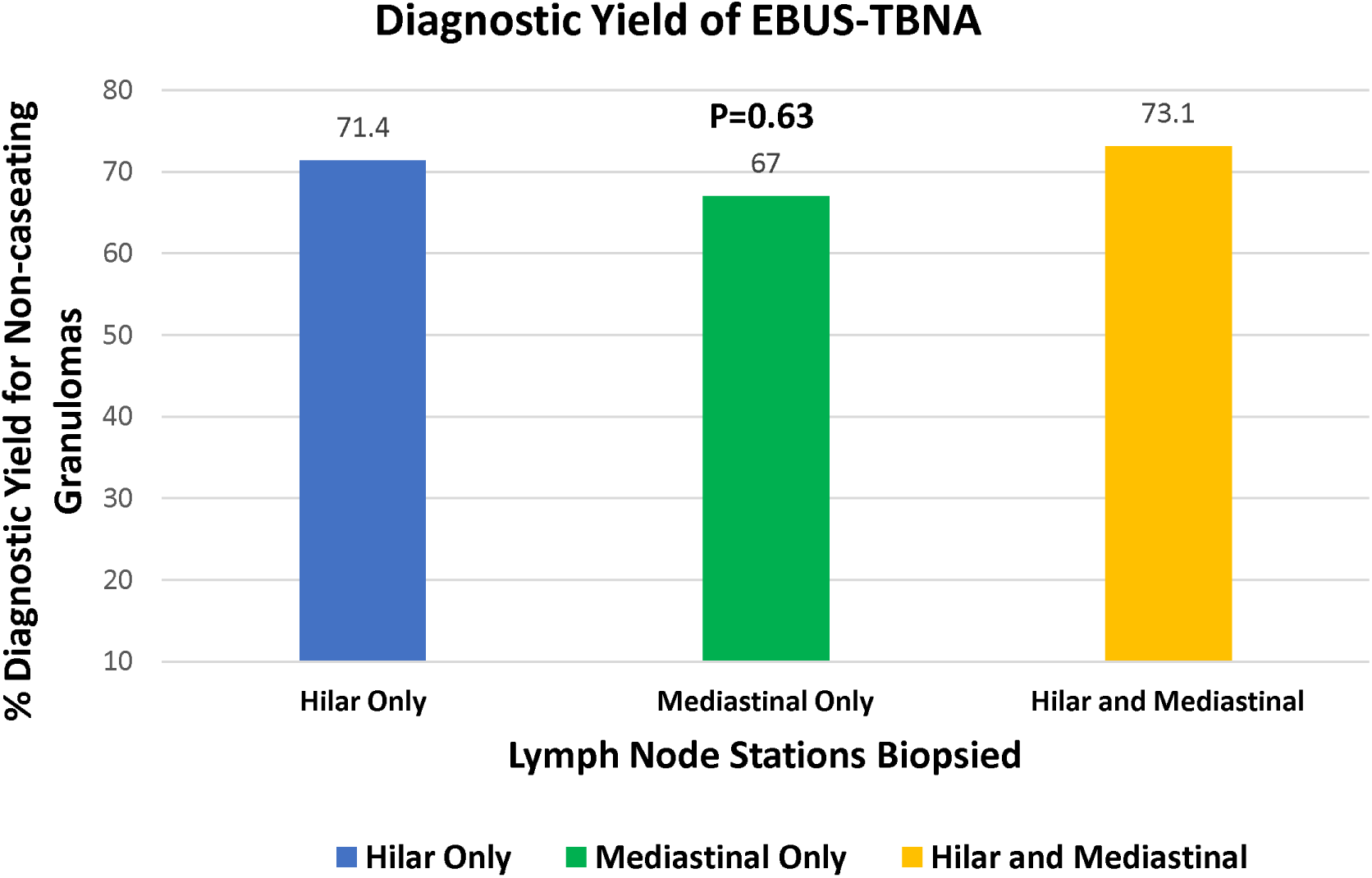
Diagnostic yield of EBUS-TBNA for non-caseating granulomas in patients with different lymph node station biopsies.

**Table 3:** Diagnostic yield of EBUS-TBNA for non-caseating granulomas based on lymph node location.

|  | Hilar Only<br>N=144 | Mediastinal Only<br>N=255 | P-value for<br>difference* |
| --- | --- | --- | --- |
| Diagnostic yield- n (%) | 89 (61.8%) | 167 (65.5%) | 0.46 |
| Sensitivity for sarcoidosis<br>(95% confidence interval) | 66.9% (58.9% - 74.9%) | 71.1% (65.3% - 76.9%) | NA** |
| Specificity for sarcoidosis | 100% | 100% | NA |

\*Chi-Square P-value
\*\* Overlap of 95% confidence interval rules out a significant difference.

**Table 4:** Diagnostic yield of EBUS-TBNA for non-caseating granulomas in patients.

|  | Hilar Nodes<br>Biopsied Only<br>(N=21) | Mediastinal<br>Nodes<br>Biopsied Only<br>(N=100) | Both Hilar and<br>Mediastinal<br>Nodes<br>Biopsied<br>(N=104) | P-value for<br>difference* |
| --- | --- | --- | --- | --- |
| Diagnostic yield- n (%) | 15 (71.4%) | 67 (67%) | 76 (73.1%) | 0.63 |

\*Chi-Square P-value

Sarcoidosis was diagnosed in 206 (91.5%) out of 225 patients in the cohort on a one-year follow-up. Definitive sarcoidosis was seen in 192 (85.3%) and probable sarcoidosis in 14 (6.2%) patients. The sensitivity for sarcoidosis of EBUS-TBNA of hilar vs. mediastinal nodes was 66.9% (95% confidence interval or CI, 58.9%-74.9%) vs. 71.1% (95% CI, 65.3%-76.9%), as shown in **Table 3**. The specificity for sarcoidosis of EBUS-TBNA of both hilar and mediastinal nodes was 100% (**Table 3**).

Univariate regression analysis showed that diagnostic yield of non-caseating granulomas was associated with number of needle passes (odds ratio, OR 1.21; 95% CI 1.02-1.44; P=0.03), age (OR 0.96; 95% CI 0.94-0.97; P <0.01) and body mass index or BMI (OR 1.05; 95% CI 1.01-1.10; P <0.01), as shown in **Table 5**. In multivariable analysis, the diagnostic yield of non-caseating granulomas was only associated with age (OR 0.96; 95% CI 0.94-0.98; P <0.01).

**Table 5:** Factors associated with diagnostic yield of non-caseating granulomas.

| Variables | Univariate Odds Ratio (95% Confidence Interval) | Univariate P value | Multivariable Odds Ratio (95% Confidence Interval) | Multivariable P value |
| --- | --- | --- | --- | --- |
| Hilar vs. mediastinal node | 0.85 (0.61, 1.20) | 0.35 | 0.96 (0.65, 1.41) | 0.82 |
| Lymph node size | 1.02 (0.99, 1.04) | 0.29 | 1.01 (0.98, 1.04) | 0.73 |
| Biopsy needle size |  | 0.11 |  | 0.35 |
| 19 vs. 22 G | 1.73 (1.00, 2.99) |  | 1.47 (0.83, 2.59) |  |
| 21 vs. 22G | 0.85 (0.26, 2.83) |  | 0.86 (0.22, 3.39) |  |
| Needle passes | 1.21 (1.02, 1.44) | 0.03 | 1.21 (0.97, 1.51) | 0.09 |
| Age* | 0.96 (0.94, 0.97) | <0.01 | 0.96 (0.94, 0.98) | <0.01 |
| Gender (female vs. male) | 0.98 (0.61, 1.58) | 0.93 | N/A |  |
| BMI** | 1.05 (1.01, 1.10) | <0.01 | 1.03 (0.99, 1.07) | 0.14 |
| Smoking status (smoker vs. non-smoker) | 1.45 (0.45, 4.68) | 0.54 | N/A |  |
| Race |  | 0.09 | N/A |  |
| Black vs. White | 1.69 (1.05, 2.73) |  |  |  |
| Others vs. White | 2.06 (0.41, 10.48) |  |  |  |

\*1-year increase
\*\* 1-BMI point increase
N/A: Excluded variables from the adjusted model due to lack of association with the outcome in univariate analysis.

## Discussion

This is one of the first studies to systematically examine the diagnostic yield of EBUS-TBNA for non-caseating granulomatous inflammation between hilar and mediastinal lymph nodes in a multiracial cohort. Although the mediastinal nodes were larger than the hilar nodes, the EBUS-TBNA diagnostic yield for either hilar or mediastinal nodes was similar. In addition, the younger the age, the higher the diagnostic yield for non-caseating granulomatous inflammation while adjusting for nodal station, node size, needle size, biopsy passes, and BMI.

In a retrospective registry, the diagnostic yield of EBUS-TBNA for non-caseating granulomas in suspected sarcoidosis patients in a propensity-matched cohort was 83.6% in the 19G group and 80.2% in the 21/22G group (P=0.60) (17). The difference in yield between hilar and mediastinal nodes was not studied. The results are similar to those of our study, as we did not find evidence of a significant difference in the diagnostic yield between different needle sizes. In a multicenter, randomized controlled trial, 185 patients with suspected sarcoidosis were randomized to EBUS-TBNA and 173 patients to endoscopic ultrasound-fine needle aspiration (EUS-FNA) (10). Granuloma detection rate was 70% for EBUS-TBNA and 68% for EUS-FNA (P=0.67). There was no comparison of diagnostic yield between hilar and mediastinal nodes, however, the diagnostic yield of EBUS-TBNA is similar to ours. In a prospective study by Sun et al. from China, 111 patients with suspected sarcoidosis underwent EBUS-TBNA (18). The diagnostic yield of EBUS-TBNA for non-caseating granulomas increased with size, stage 1 sarcoidosis (compared to stage 2), and ≥2 needle passes (compared to one pass). EBUS-TBNA showed granulomas in 79 out of 103 (76.6%) hilar 148 out of 181 (81.7%) mediastinal lymph nodes, P=0.54. Our study extends the literature as the cohort is multiracial, and the details of nodal size, needle size, and number of needle passes are compared between hilar and mediastinal nodes. Although younger patients had a higher diagnostic yield of non-caseating granulomas, node size, and needle passes were not associated with a higher yield. We speculate that the needle passes were not associated with a higher diagnostic yield in our study as the median number of needle passes was four per node, which is already above the range of 3-4 needle passes where the diagnostic yield of EBUS-TBNA reached a plateau in Sun’s and others studies (18, 19). In a meta-analysis, including 14 studies and 2097 patients, the EBUS-TBNA yield varied from 33% to 100%, but the mean pooled diagnostic yield was 0.79 (standard deviation, 0.24) (20). In another meta-analysis, a similar pooled diagnostic yield of 79% (95% CI, 71-86%) was reported, and ROSE did not affect the diagnostic outcome (21). The diagnostic yield of EBUS-TBNA in our cohort is similar. The sensitivity and specificity of EBUS-TBNA for sarcoidosis in our cohort are similar to what is reported in other retrospective studies.(20)

We found that odds of finding non-caseating granulomas increased with younger age in univariate and multivariate analyses. Sarcoidosis in the US is generally seen at younger age, though there has been a shift to older age in the recent studies (2, 5, 6). The median age (IQR) of our cohort was 54 years (43–63), similar to the published literature (2, 5, 21). A study evaluated the association of age with cytological diagnosis of sarcoidosis in 89 patients who underwent EBUS-TBNA (22). Younger age was an independent predictor of cytological diagnosis of non-caseating granulomas in univariate and multivariable regression analysis, similar to the findings of our study.

The strengths of this study include a detailed assessment of EBUS-TBNA yield of hilar and mediastinal nodes in a large, multiracial cohort of patients with sarcoidosis. The data included details of hilar vs. mediastinal biopsy location, lymph node size, different needle sizes, and number of needle passes. Our study is limited by its retrospective, single center design. Because of the utilization of ROSE, not all patients underwent both hilar and mediastinal node biopsies, and the procedure was stopped if a hilar or mediastinal node aspirate showed non-caseating granulomas. However, 104 out of 224 patients (46.4%) underwent biopsies of both hilar and mediastinal stations. Future prospective, multi-center studies are warranted to assess this further.

### Conclusions

In conclusion, we did not find a statistically significant difference in the diagnostic yield of EBUS-TBNA between hilar and mediastinal lymph nodes. If clinically warranted, we recommend patients with suspected sarcoidosis should undergo biopsy of both hilar and mediastinal stations, especially if ROSE is not utilized.

## Data Availability

All data produced in the present study are available upon reasonable request to the authors.

## Acknowledgements

We acknowledge the struggle and persistence of our patients with sarcoidosis, their families, and healthcare providers managing them.

## Reporting Checklist

We present this manuscript in accordance with the STARD reporting checklist.

## Data Availability Statement

The data that support the findings of this study are available from the corresponding author upon reasonable request.

