## Supplemental Table 1 for "EBUS Diagnostic Yield for Sarcoidosis in Hilar vs. Mediastinal Lymph Nodes"

**Table S1: Results of transbronchial and endobronchial biopsies.**

| Transbronchial biopsy site- n (%)  RUL  RML  RLL  LUL  Lingula  LLL  RUL+RLL  RUL+RML  Not performed | 20 (8.9)  4 (1.8)  38 (16.9)  3 (1.3)  1 (0.4)  7 (3.1)  1 (0.4)  1 (0.4)  150 (66.7) |
| --- | --- |
| Transbronchial biopsy results – median (%)  Non-caseating granulomas  Cancer  Alveolar tissue  Other  Not performed | 31 (13.8)  1 (0.4)  33 (14.7)  10 (4.4)  150 (66.7) |
| Endobronchial biopsy site - n (%)  Trachea  Right mainstem  Bronchus intermedius  RUL  RML  RLL  Left mainstem  LUL  Lingula  LLL  Trachea+RUL Trachea+RML  RML+LUL+Lingula  Random  Not performed | 12 (5.3)  4 (1.8)  1 (0.4)  11 (4.9)  3 (1.3)  7 (3.1)  4 (1.8)  5 (2.2)  0  0  1 (0.4)  1 (0.4)  1 (0.4)  1 (0.4)  174 (77.3) |
| Endobronchial biopsy results- n (%)  Non-caseating granulomas  Cancer  Bronchial epithelium  Not performed | 29 (12.9)  2 (0.9)  20 (8.9)  174 (77.3) |
